# Spatial patterns of excess mortality in the first year of the COVID-19 pandemic in Germany

**DOI:** 10.1101/2022.03.10.22272221

**Authors:** Thomas Wieland

**Affiliations:** Karlsruhe Institute of Technology

## Abstract

In order to quantify the impact of the SARS-CoV-2/COVID-19 pandemic, several studies have estimated excess mortality rather than infections or COVID-19-related deaths. The current study investigates excess mortality in Germany in 2020 at a small-scale spatial level (400 counties) and under consideration of demographic changes. Mortality is operationalized using standardized mortality ratios (SMRs), visualized on maps, and analyzed descriptively. Regional mortality and COVID-19-related morbidity are tested for spatial dependence by the Moran’s I index. It is, furthermore, tested whether all-cause mortality is associated with COVID-19-related morbidity by correlation coefficients. Excess mortality only occurrs in a minority of counties. There are large regional disparities of all-cause mortality and COVID-19-related morbidity. In older age groups, both indicators show spatial dependence. (Excess) mortality in older age groups is impacted by COVID-19, but this association is not found for young and middle age groups.

## 1 Introduction

Since the emergence of the *Severe Acute Respiratory Syndrome Coronavirus-2* (*SARS-CoV-2*) and the *Coronavirus disease 2019* (*COVID-19*), which is the respiratory illness caused by SARS-CoV-2, there has been a discussion about the overall societal effects of the pandemic. These impacts can be, for example, an overwhelming of the healthcare system (due to many serious illnesses at the same time) or, in the worst case, deaths attributed to COVID-19. Estimates of the infection fatality rate (IFR) of SARS-CoV-2/COVID-19 from epidemiological studies range between 0.00% and 1.54% with a median value of 0.23%, with the risk increasing drastically with age (Ioannidis 2021). Based on a literature review, a German study defines all people of 65 years and older as the COVID “risk group”, with a number of previous illnesses (e.g. heart disease, obesity) playing an important role in the probability of a severe course (Rommel et al. 2021).

To account for the “burden of disease” of the pandemic, many studies look at excess mortality instead of using data on confirmed infections and deaths because this data is often subject to uncertainties (e.g., variations in testing, definition of a “COVID-19 death”). Excess mortality describes an increased mortality compared to an expected value. Analysis of excess mortality allows mortality from COVID-19 to be quantified by comparing actual deaths with expected deaths that would have occurred without the pandemic (Kowall et al. 2021, Stang et al. 2020). For the first year of the pandemic, 2020, the Federal Office of Statistics of Germany estimates a number of 71,000 excess deaths (Statistisches Bundesamt 2021b). Kowall et al. (2021) and Stang et al. (2020) argue that the analysis of mortality over time must incorporate demographic changes within the society, as a disproportionate increase of older age groups *must* lead to higher mortality. In Germany, the population increased from 82.2 millions in 2016 to 83.2 millions in 2020 (+1.21%). In the same time, inhabitants of 80 years and older increased from 4.7 millions to 5.7 millions (+20.13%) (Statistische Ämter des Bundes und der Länder 2022a). Whilst accouting for demographic changes, Kowall et al. (2021) analysed excess mortality in Germany, Sweden and Spain, and can not find excess mortality in Germany in 2020.

From the perspective of spatial epidemiology and health geography, the spread of an infectious disease must be regarded as a spatial phenomenon (Elliott, Wartenberg 2004). The spread of a virus (or, more general, pathogen) is a spatial diffusion process, with regional differences in the transmission *within* and *between* regions (Charu et al. 2017, Viboud et al. 2006). Infection waves can be very asynchronous between regions of the regarded country and can also vary greatly in severity. This is due to population heterogeneity, which includes interpersonal differences in contact networks and “super-spreading events” at the local level, with both of them resulting in strong regional variations in timing and extent of outbreaks (Chowell et al. 2015, Thomas et al. 2020). Transmission between regions is driven by spatial interactions. There are spillovers of transmission due to human mobility across administrative borders such as commuting (Charaudeau et al. 2014, Charu et al. 2017, Dalziel et al. 2013, Findlater, Bogoch 2018, Viboud et al. 2006). Studies have found both regional disparities and spatial dependence of COVID-19 cases and deaths for the first pandemic wave in spring 2020 (Bourdin et al. 2021, Saffary et al. 2020, Wang et al. 2021, Wieland 2020). It is, thus, to be expected a) that there are also regional disparities in (excess) mortality, and b) that (excess) mortality levels in nearby regions are more similar than with respect to more distant regions.

The current study investigates excess mortality in Germany in 2020. According to the above conditions, mortality is analysed a) at a small-scale spatial level, and b) under consideration of demographic changes in Germany. The analysis is conducted at the level of counties (in German: *Landkreise, N* = 400) which is the second smallest spatial unit for which official statistical data is available in Germany, and the smallest unit for which COVID-19 cases and deaths are available. The first aim of the present study is the descriptive analysis and map visualization of excess mortality at the county level for Germany in 2020. Beyond this, the study tests regional mortality and COVID-19-related morbidity for spatial dependence. In the last step, it is investigated whether regional mortality can be attributed to COVID-19, more precisely, it is tested whether all-cause mortality is associated with COVID-19-related morbidity.

The data used for the analysis and the related statistical methods are presented in the next section (section 2). The results are shown in section 3 and discussed in section 4. In section 5, conclusions and limitations of the study are presented.

## 2 Material and methods

### 2.1 Data

Data on regional all-cause deaths was retrieved from the *Regionaldatenbank Deutschland*, which provides official statistics at the subnational level (16 states, 400 counties, approx. 11,000 municipalities). Table 12613-93-01-4 was used, which contains total all-cause deaths by age group at the county level (Statistische Ämter des Bundes und der Länder 2022b). The county dataset does not include mortality data disaggregated by age group *and* sex *and* time (monthly or weekly). Thus, the current analysis is conducted at the cumulative level (whole year) and for age groups. In some cases, total deaths for a specific age group in a specific county are not available because the numerical value is unknown or not to be disclosed (due to data protection laws). This only applies for younger age groups (*<*35) in some counties with a small population size. These counties were excluded in the subsequent analysis of the respective age groups.

Regional population sizes by age group was extracted from the same service using table 12411-09-01-4 (Statistische Ämter des Bundes und der Länder 2022a). As the subsequent analysis relates to mortality during a given year, the population in year *t* is regarded as the population at December 31st of year *t* -1.

The total number of COVID-19-related deaths at the county level was extracted from the COVID-19 case dataset, which is provided by the German *Robert Koch Institut* (*RKI*), Germany’s governmental public health institute. This dataset includes all case reports including information on age group, sex, place of residence (county), date of confirmation, and, in some cases, date of onset of symptoms, as well as the information whether it is a case of death or not (Robert Koch Institut 2022c). Note that this dataset includes people who died directly from COVID-19 (“died from”), as well as deceased people who were infected with SARS-CoV-2 and for whom it cannot be conclusively proven what the cause of death was (“died with”), especially in the case of serious pre-existing conditions. The decision as to what is classified as a COVID-19 death is up to the respective regional health department (Robert Koch Institut 2022a). This dataset does not provide information about the date of death, and available statistics about weekly COVID-19-related deaths are not published at the county level (Robert Koch Institut 2022d). Thus, COVID-19 deaths had to be extracted based on the date of confirmation by comparing the monthly confirmed deaths with the sum of COVID-19 deaths on condition of a specific confirmation date. The best match was achieved when using a confirmation date up to 2020/12/21 with 42,063 deaths, which is very close to the official numerical value of 41,648 COVID-19-related deaths in the dataset containing deaths by month.

Boundaries of German counties were retrieved from the county dataset (shapefile) provided by the RKI (Robert Koch Institut 2022b).

The data sources had to be harmonized due to different age categories. Mortality and population data were adjusted to the age groups in the RKI dataset (*<*5, 5-14, 15-34, 35-59, 60-79, 80+). As there have been few COVID-19-related deaths in the age groups below 35, the first three age groups (*<*5, 5-14, 15-34) were aggregated, which leads to four age groups in the subsequent analysis (0-34, 35-59, 60-79, and 80+). County boundaries and COVID-19 death numbers had to be harmonized as well because, in the RKI dataset, the capital Berlin is divided into 11 districts. By contrast, in the regional mortality dataset, Berlin is processed as a whole. Thus, COVID-19 deaths had to be summed up over whole Berlin.

### 2.2 Statistical analysis

Following previous studies on excess mortality (Kowall et al. 2021, Morfeld et al. 2021, Stang et al. 2020), *standardized mortality ratios* (*SMR*s) were calculated for each age group (0-34, 35-59, 60-79, and 80+) and county (*N* = 400). SMRs compare observed mortality in a given time with expected mortality, with the latter being derived from previous mortality in a reference period. Following Kowall et al. (2021), to account for demographic changes over time, the observed and expected mortality was calculated from age-specific *mortality rates* instead of the age-specific number of total deaths. The mortality rate (sometimes referred to as *crude death rate*) of age group *a* in year *t* and region *r* is:

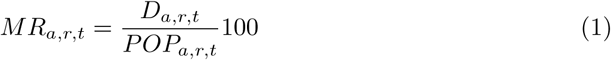

where *D*_*a,r,t*_ is the number of all-cause deaths in age group *a* in year *t* and region *r*, and *POP*_*a,r,t*_ is the population size in age group *a* at year *t* and region *r*.

Here, the expected mortality rate for 2020 is defined as the median of the mortality rates in the reference period. Following previous studies on COVID-related excess mortality in Germany (Kowall et al. 2021, Morfeld et al. 2021, Stang et al. 2020, Statistisches Bundesamt 2021b), the reference period is 2016-2019. The standardized mortality ratio for age group *a* in year *t* and region *r* is defined as:

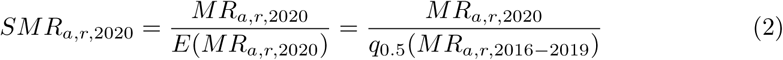

COVID-related morbidity was calculated as the cumulative number of COVID-19 fatalities in age group *a* in region *r* at time *t* relative to the corresponding population:

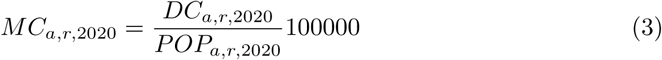

where *DC*_*a,r*,2020_ is the number of COVID-19-related deaths in age group *a* in 2020 and region *r*, and *POP*_*a,r*,2020_ is the population size in age group *a* at 2020 in region *r*.

To account for spatial dependence, the age group specific indicators for excess mortality and COVID fatalities were tested for spatial autocorrelation using *Moran’s I* coefficient. This index measures overall spatial autocorrelation in terms of a regression of regional values and values of nearby spatial units. The expected value of *I* depends on the number of regions: *E*(*I*) = −1*/*(*N* − 1). The test for statistical significance investigates whether the observed value of *I* is significantly greater than the expected value (Bivand, Wong 2018, Griffith 2009). This analysis requires a weighting matrix for the definition of spatial proximity of the regarded spatial units. Here, the weighting includes all neighboring counties, which means that the weighting for county *r* with respect to another county is equal to one if they are adjacent and equal to zero if not.

To quantifiy relationships between regional mortality and COVID morbidity, the indicators were tested for statistical dependence by Pearson correlation coefficients (*r*).

As one cannot expect that all indicators of mortality and morbidity are normally distributed, they were transformed by natural logarithm for the spatial autocorrelation and correlation analysis. If a given variable contains values equal to zero (which may occur with respect to age-specific morbidity, not mortality), a small constant was added to enable the analyses (ln(*x* + 0.01)). In all significance tests, the significance level was set to 95% (*p <* 0.05). All analyses were conducted in *R* (R Core Team 2021), while using the packages *spdep* (Bivand, Wong 2018) and *corrplot* (Wei, Simko 2021).

## 3 Results

Standardized mortality ratios (SMR) for all four age groups at the county level are shown in the maps in figure 1. In the maps, the SMRs are classified in steps of 0.05 (5%) with values above one reveal that the observed mortality is above the expected mortality (excess mortality). As there is no (complete) data for 72 counties with respect to the first age group (0-34), all subsequent statistics were calculated for the remaining 328 counties. Figure 2 presents regional COVID-19 morbidity based on the *RKI* data and classified by quantiles (except for the first age group as there are no COVID-19 fatalities in the majority of counties). In figure 3, the correlations between the aforementioned indicators are visualized. Non-significant Pearson correlation coefficents (*p >* 0.05) are crossed out.

**Figure 1:**
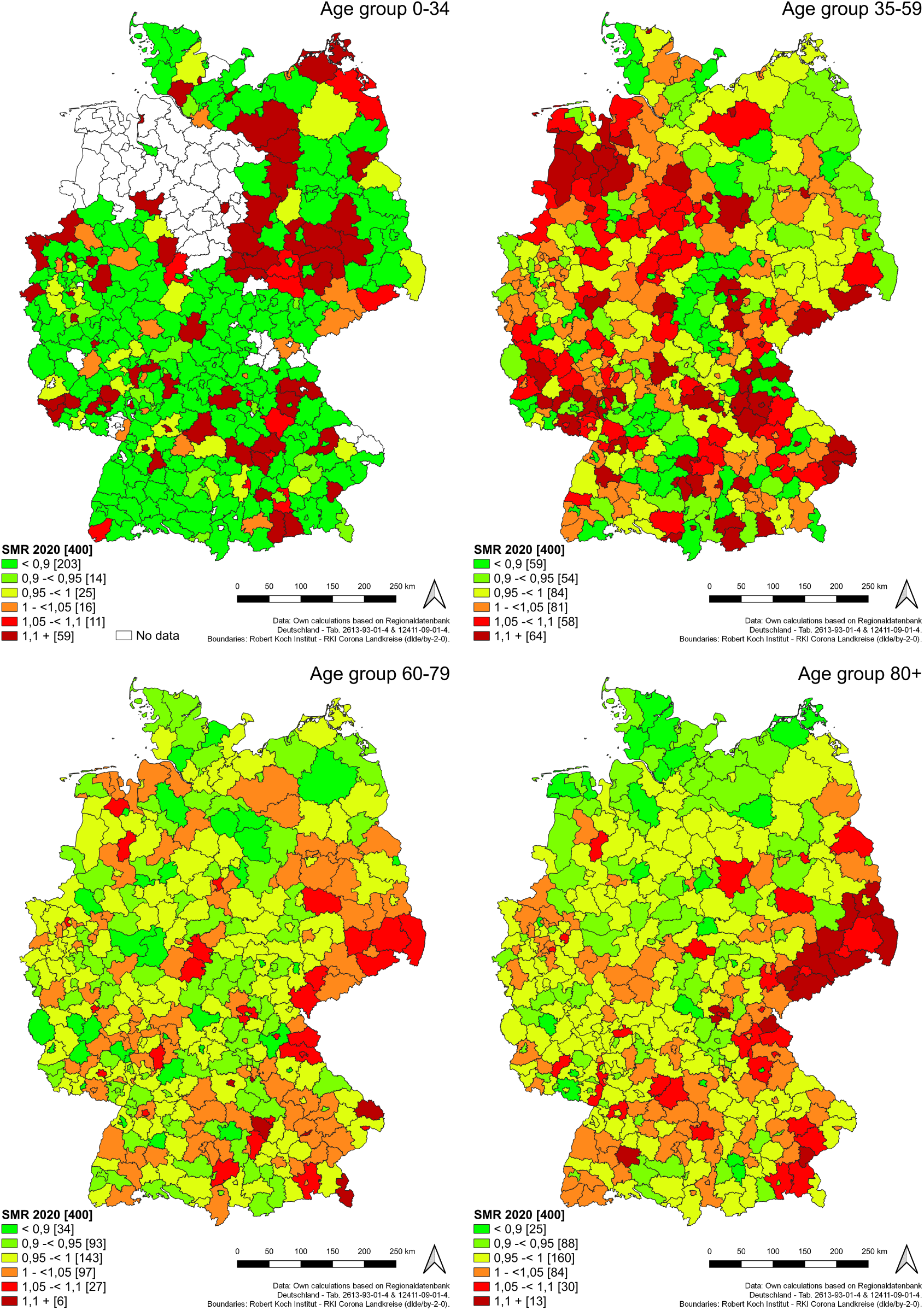
Standardized mortality ratios (SMRs) at the county level 2020

**Figure 2:**
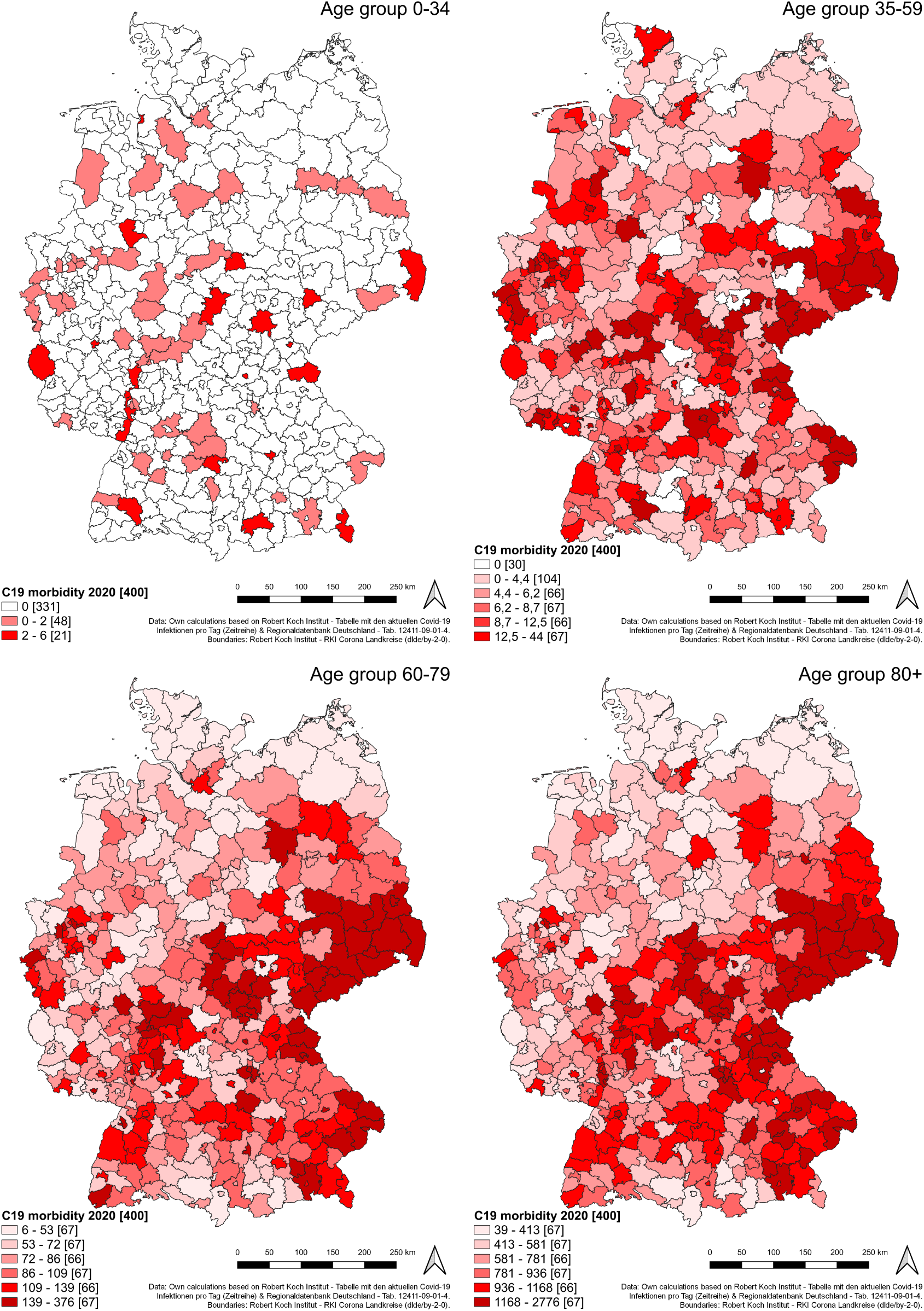
COVID-19-related morbidity at the county level 2020

**Figure 3:**
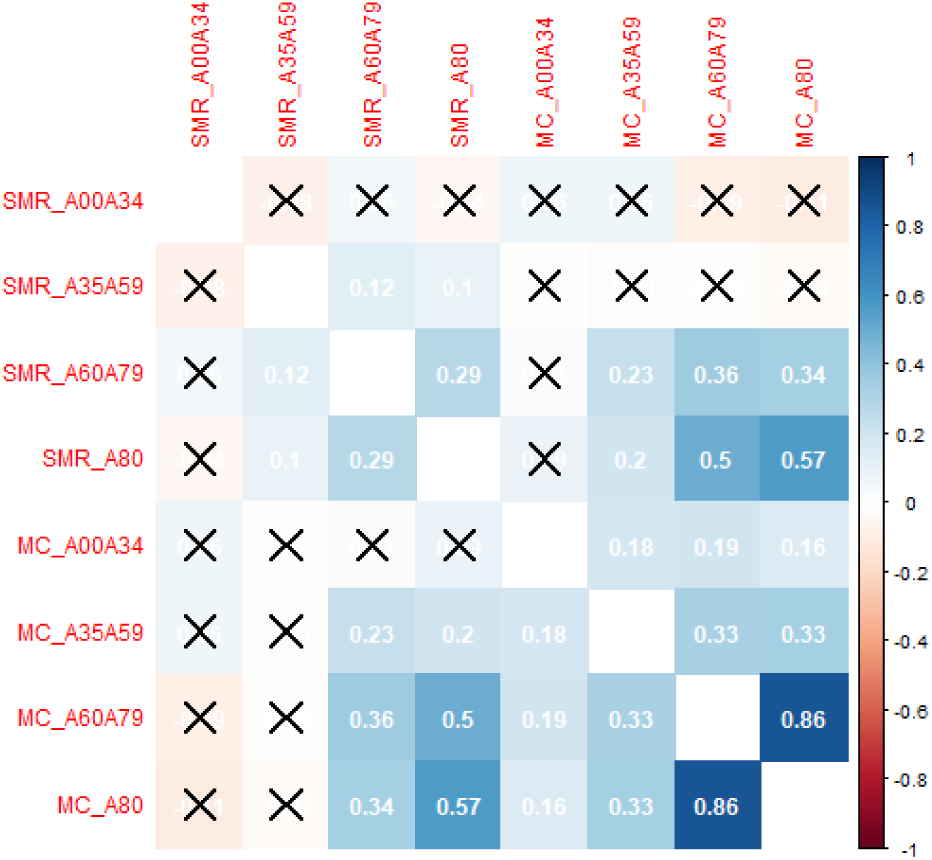
Pearson correlation coefficients for SMRs and COVID-19 morbidity at the county level

The SMRs for the age group 0-34 (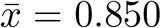, sd = 0.301, *q*_0.5_ = 0.869) are distributed as follows. In the vast majority of counties (242; 73.78%), the observed mortality in 2020 in below the expected value (*SMR <* 1), while there is excess mortality (*SMR >* 1) in the remaining 86 counties (26.22%). In 59 counties (17.99%), excess mortality is equal to 10% or higher (*SMR* ≥ 1,1). Because of the lack of data which is obviously unevenly distributed over space, no Moran’s I coefficient was estimated for the first age group. For age group 35-59, the coefficient for spatial autocorrelation, Moran’s I, is equal to *I* = 0.020 (*p* = 0.25), and, thus, not significant above the expected value of *E*(*I*) = -0.003 (which is equal for all age groups due to the same number of spatial units). Taking a look at the values of all counties (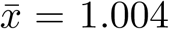, sd = 0.102, *q*_0.5_ = 0.999) shows that 203 counties (50.75%) reach values of *SMR >* 1, which means that there is excess mortality in a narrow majority of counties for this age group. However, there is no signíficant spatial dependence of mortality. This is different from the SMR of the third age group, 60-79 (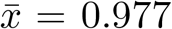, sd = 0.053, *q*_0.5_ = 0.975). In 132 counties (33.00%), the SMRs are above one, with an excess mortality of 10% or above (*SMR* ≥ 1,1) in six counties (1.5%). The estimate of spatial dependence is *I* = 0.085, which is significantly higher than the expected value (*p <* 0.01). This result is similar with respect to the age group of 80 and above (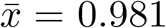, sd = 0.055, *q*_0.5_ = 0.977). Here, Moran’s I has the value of *I* = 0.350 (*p <* 0.01). SMR values above one are found in the 127 of 400 counties (31.75%), which means that excess mortality occurs in a minority of German counties. In 13 counties (3.25%), excess mortality reaches 10% or more. All in all, excess mortality can be identified in a minority of counties, except for one age group. Mortality is spatially autocorrelated for elder age groups (60-79, 80+) with a geographical emphasis in the east and southeast of Germany (states Saxony and Bavaria), but not for younger age groups.

Taking a look at the maps in figure 2, there are obvious spatial differences in the cumulative COVID-19 morbidity (COVID-19 deaths per 100,000 pop) in 2020. In the age group 0-34 (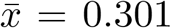, sd = 0.817, *q*_0.5_ = 0.000), there are 331 counties without COVID-19-related deaths in 2020. The indicator for spatial dependence, Moran’s I, is equal to *I* = 0.0005, and not significant (*p* = 0.46). In contrast, COVID morbidity in the age group of 30-59 (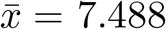, sd = 5.785, *q*_0.5_ = 6.240) is spatially autocorrelated (*I* = 0.132, *p <* 0.01). The same can be found for the age groups 60-79 (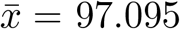, sd = 52.865, *q*_0.5_ = 86.438) with *I* = 0.503 (*p <* 0.01) and 80+ (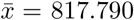, sd = 444.788, *q*_0.5_ = 781.434) with *I* = 0.519 (*p <* 0.01). COVID-19-related morbidity is, thus, spatially autocorrelated with respect to the age groups 30-59, 60-70, and 80+. Spatial clustering of morbidity can be identified especially in the east and southeast of Germany.

It is, thus, not surprising that COVID-19-related morbidity in the elder age groups (60-79, 80+) correlates strongly and positively, with a Pearson correlation coefficient of *r* = 0.86 (*p <* 0.01). Regional COVID-19 morbidity in the other age groups show weak to moderate positive correlations (*r* = 0.16 to *r* = 0.33, *p <* 0.01) (see figure 3). Whilst the SMR and morbidity values are not significantly correlated in younger age groups (0-34, 35-59), there are significant correlations between SMRs and COVID-19 morbidity for the age groups 60-79 (*r* = 0.36, *p <* 0.01), and 80+ (*r* = 0.57, *p <* 0.01). The latter correlation coefficient corresponds to *r*^2^ = 0.32, which means that 32% of the variance in regional mortality is explained by regional COVID-19 morbidity in the age group of 80+. Thus, there is, at least to a certain degree, an association between regional COVID-19 morbidity and regional all-cause mortality in elder age groups, but this correlation can not be confirmed for younger age groups.

## 4 Discussion

Whilst taking into account demographic changes, Kowall et al. (2021) have found that there has been no excess mortality at the national level in Germany in the first pandemic year. In principle, the current results confirm these findings, as regional excess mortality occurred only in a minority of counties, with most of them showing mortality values below the expected mortality instead. These findings are clear with respect to young (0-34) and older age groups (60-70, 80+), but there is a balance of counties with and without excess mortality for the middle age group (35-59).

However, this result does not rule out excess mortality due to COVID-19 *per se*. There are regions with mortality clearly above the expected value in all age groups, and it is, thus, an interesting question whether this may be attributed to the pandemic. Both excess mortality and COVID-19 morbidity in the older age groups (60-79, 80+) tend to be spatially clustered, which was shown in the maps. Especially counties in the east of Saxony and Bavaria have experienced excess mortality of 10% or more in the age group 80+. At least a part of regional excess mortality in the older groups (especially 80+) is statistically associated with COVID-19 morbidity, as shown in the correlation analysis. Thus, it is likely that COVID-19 has contributed substantially to excess mortality in the age-specific COVID-19 “risk group”. In contrast, regional (excess) mortality of people below 60 years can *not* be explained by COVID-19, as there is (a) no such spatial pattern in mortality and (b) no correlation between COVID deaths and all-cause mortality.

The analysis of spatial autocorrelation has shown spatial dependence of both regional mortality and COVID-19 morbidity, at least for the older age groups. This can be explained by a spatially clustered occurrence of infections in combination with interregional virus transmission due to interregional mobility. As there are more spatial interactions between nearby regions, also infection levels of nearby regions are more similar than distant ones. The influence of mobility (especially commuting) on virus transmission was outlined several times for infectious diseases such as influenza (Charaudeau et al. 2014, Charu et al. 2017, Dalziel et al. 2013, Viboud et al. 2006). With respect to SARS-CoV-2, Mitze, Kosfeld (2021) outline the enhancing effect of commuting to work on regional infections in the first pandemic wave in Germany. Regarding the same time period, Wieland (2020) shows that, all other things being equal, growth rates of SARS-CoV-2 infections in German counties increase with increasing intensity of commuting. Bourdin et al. (2021) find that the regions most affected by COVID-19 in the first wave in Italy are also those with the highest level of connectivitiy to the rest of the world. The present results towards regional mortality and COVID-19 morbidity are in line with studies which have found spatial dependence of SARS-CoV-2 infections and/or COVID-19 deaths at the small-scale level in the initial phase of the pandemic, such as in Italy (Bourdin et al. 2021), USA (Saffary et al. 2020), Germany (Wieland 2020), and China (Wang et al. 2021).

The occurring of spatial clusters (“hotspots”) of (COVID-associated) excess mortality and COVID-19 fatalities can be caused by both interregional and intraregional disease transmission. Regional disparities of infections and deaths have been confirmed for many epidemics, regardless of the pathogen. The main reason is population heterogeneity, which means that individuals do not have the same probability of being infected and/or infecting others, especially because of heterogeneous networks of social contacts. Network connectivity may differ between regions. Furthermore, there are “super-spreading events” at the local level, at which many people get infected at once. These reasons lead to spatial heterogeinity in infectious disease epidemics with strong differences in regional infections or deaths (Chowell et al. 2015, Thomas et al. 2020). The study cannot examine the reasons for the hotspots found in the analysis directly, but could provide possible interpretations. Some regional hotspots in the first wave of infections are attributed to specific super-spreading events, such as indoor public large-scale events (Brandl et al. 2021, Streeck et al. 2020). There have also been outbreaks in nursing homes across Germany in 2020, resulting in many COVID-19 related deaths (Kohl et al. 2021, Wieland 2020).

Clusters of excess mortality and COVID-19 morbidity were especially found in the east and southeast of Germany. As these counties are located at the Czech and Polish border, commuting from these countries might explain a higher level of infections, since their border regions were themselves hotspots at times. Many of these cross-border commuters work in hospitals, nursing homes and home care, and are, thus, in contact with people of the age-specific risk group (Ärzteblatt 2020a,b, Berliner Morgenpost 2021). It is therefore possible that this could explain the increased mortality in these regions. However, this cannot be proven, as most chains of infection are not traceable. Such cross-border transmission has been confirmed for the first care home outbreak in the Netherlands, which was traced back to mobility from Germany (Mitch van Hensbergen 2021).

There is a possible indication of population heterogeneity in the results. Except for the age groups 60-79 and 80+, regional COVID-19 morbidity shows only weak to moderate correlations between the four regarded age groups. This means that the amount of COVID-related deaths within one age group in a given region is hardly associated with the amount of COVID-related deaths within another age group in the same region. The further apart the age groups are, the smaller the correlations are. This might be an indication that virus transmission between people of different age groups within the regions was rather low. These results may be explained by a strong *age assortativity*, which means that individuals tend to have more social contacts with people of similar age. This fact is well-known in epidemiology and proven by many studies on social mixing patterns (Leung et al. 2017, Mossong et al. 2008, Read et al. 2014).

## 5 Conclusions and limitations

In the present study, mortality for Germany in 2020 at the county level (*N* = 400 spatial units) was estimated, whilst taking into account demographic changes from 2016 to 2020. Regional excess mortality was tested for spatial dependence and for correlation with regional COVID-19-related morbidity. It was shown that excess mortality only occurred in a minority of counties, which is consistent with results of previous studies on the national level. There are large regional disparities of all-cause mortality and COVID-19-related morbidity. In older age groups, both indicators show spatial dependence. (Excess) mortality in older age groups is impacted by COVID-19, but this association is not found for young and middle age groups. The present results lead to two main conclusions:

- Epidemics and pandemics have to be regarded as spatial phenomenons. The spread of a pathogen or disease is a spatial diffusion process with strong regional disparities and spatial dependence. Transmission varies within and between regions, which results from population heterogeneity and interregional mobility. Any kind of analysis or forecast should be conducted at a small-scale spatial level. Indicators such as incidence, deaths or excess mortality are not very meaningful at national or at a roughly delineated sub-national level such as federal states.
- COVID-19 in 2020 has had the potential to increase mortality in older age groups well above the expected mortality. In contrast, this effect has not been determined for people of age groups below 60 years. Consequently, *if* preventing COVID-related deaths is defined as the primary goal of governmental interventions, then protective measures (*Nonpharmaceutical interventions, NPI*) should be (or should have been) aimed in particular at the COVID-19 risk group, especially older people. Thus, prioritizing vaccination (which started at the end of December 2020) by age and protective measures for nursing homes can be considered useful.

The analysis faces some limitations with respect to the interpretation of the results:

- Although spatial clusters and spatial dependence of excess mortality and COVID-19 deaths were revealed, the study can not identify *why* these regional disparities occur. Possible explanations that have been formulated are just conjectures rather than evidence. This is because the context of infection is unknown for the most confirmed SARS-CoV-2 infections and COVID-19 deaths in Germany. Hotspots at a regional level are based to a large extent on region-specific singularities, and it will be an important task for future studies in health geography and spatial epidemiology to find out why certain regions have been hit so hard by COVID-19 and others not.
- The interpretation of (excess) mortality in 2020 is difficult due to specific circumstances in Germany in this year. For example, the wave of seasonal influenza in the winter season 2019/2020 was extraordinarily mild and stopped at the beginning of March 2020 (Buchholz et al. 2020). Additionally, the reduction of social contacts and mobility might have had positive side effects in terms of a reduction of all-cause mortality. For example, in 2020, traffic accident fatalities were 10.6% lower than in 2019 (Statistisches Bundesamt 2021a). There might have also been negative side effects (“collateral damages”). For example, Kortüm et al. (2020) have found excess mortality in the German county Waldshut in April 2020, with only 55% of this can be attributed to COVID-19 deaths. As the number of patients in emergency care declined strongly during this period, the authors assume that the remaining 45% can be attributed to missed treatments of other severe diseases.
- There is the question whether have impacted (excess) mortality and COVID-19 morbidity. Previous studies on excess mortality in Germany have stated that they cannot assess the effect of these measures due to potential confounding factors (Morfeld et al. 2021, Stang et al. 2020). As there are large regional disparities of both mortality and COVID-related morbidity, it remains unclear whether and, if so, by how much COVID-19 deaths were reduced by the interventions. This question has also been raised in an international context. In a country-level analysis, Chaudhry et al. (2020) can not find significant influences of border closures, full lockdowns, and test volume on COVID-19-related deaths per million people. For 2020, Kowall et al. (2021) find no excess mortality in Germany, little excess mortality in Sweden, but strong excess mortality in Spain.
- It is important for interpretation to remember that the spatial scale has a substantial impact on the result of such an analysis. Like with many other variables, variance of mortality and disease fatalities increases with the resolution of the spatial units, and, thus, the more small-scale the analysis is, the more heterogeneous is the overall picture. This is a part of the *Modifiable Areal Unit Problem* (*MAUP*), which is well-known in the spatial sciences (Elliott, Wartenberg 2004, Manley 2014). Moreover, regional disparities in all-cause mortality can be attributed to many reasons (e.g., health-related behaviour, living conditions, local climate) and have been determined in Germany before (Robert Koch Institut 2011). Thus, an explanation of spatial mortality patterns *in general* is outside the scope of this study.

Apart from the interpretation, the study also has methodological limitations:

- In contrast to previous studies, the present analysis does not investigate mortality over time, as the data provided by the German *Regionaldatenbank* is limited at this spatial scale (counties). E.g., there is monthly (not weekly) data, but not disaggregated by age groups. Apart from that, it is questionable whether the results of such an analysis with 400 spatial units would still be comprehensible. One has to keep in mind that the present results relate to *cumulative* mortality in one year, which does not rule out excess mortality within a specific time period.
- Mortality data for some age groups is incomplete at the county level, which has lead to leaving out these counties from the mortality analysis for the first age group.
- In fact, it is unknown how many people died by (or with) COVID-19 at the county level *within a given time period*. Thus, county-level COVID deaths had to be estimated based on the best match for a specific confirmation date. This can lead to inaccuracies in determining regional COVID deaths.

Finally, it must be reiterated that the analysis conducted here is aimed at the societal impact of the COVID-19 pandemic (like any other study towards excess mortality), not the risk to the individual. The presence or absence of excess mortality does not say anything about how dangerous the infection with a virus or the disease it causes is for a person. Once mortality data for 2021 are available at the county level, this analysis should be repeated for the second year of the pandemic. The general conditions have changed significantly because vaccination started at the end of December 2020, where the elderly were prioritized. Furthermore, a SARS-CoV-2 testing strategy in nursing homes was established around the same time. Additionally, new virus variants emerged in 2021, which may have influenced COVID-related morbidity as well.

## Data Availability

All data produced in the present study are available upon reasonable request to the authors

